# Quantitative cone contrast threshold testing in patients with differing pathophysiological mechanisms causing retinal diseases

**DOI:** 10.1101/2020.11.10.20228619

**Authors:** Kayla M. White, Caroline Frambach, John Doan, Urmi V. Mehta, Itamar Livnat, Clara Yuh, Anton M. Palma, Kimberly A. Jameson, M. Cristina Kenney, Mitul C. Mehta, Chantal J. Boisvert, Wade R. Crow, Andrew W. Browne

## Abstract

**Purpose:** Cone contrast threshold testing (CCT) provides quantitative measurements of color and contrast function to reveal changes in vision quality that is not a standard endpoint in clinical trials. We utilize CCT to measure visual function in patients with multiple sclerosis (MS), age-related macular degeneration (AMD), epiretinal membrane (ERM), and retinal vein occlusion (RVO).

**Methods:** Retrospective data was gathered from 268 patients of the Gavin Herbert Eye Institute. Subjects included 17 patients with MS, 45 patients with AMD, 41 patients with ERM, 11 patients with RVO, and 123 age and visual acuity-matched healthy controls. Patients underwent the primary measurement outcome, CCT testing, as well as Sloan visual acuity test and spectral domain optical coherence tomography during normal care.

**Results:** Color and contrast deficits were present in MS patients regardless of history of optic neuritis. AMD with intermediate or worse disease demonstrated reduced CCT scores. All 3 stages of ERM demonstrated cone contrast deficits. Despite restoration of visual acuity, RVO-affected eyes demonstrated poorer CCT performance than unaffected fellow eyes.

**Conclusions:** CCT demonstrates color and contrast deficits for multiple retinal diseases with differing pathophysiology. Further prospective studies of CCT in other disease states and with larger samples sizes is warranted.

**Brief Summary Statement:** In a retrospective analysis of 268 adults, cone contrast threshold testing (CCT) demonstrates patterns of visual function deficits in multiple sclerosis and age-related macular degeneration and similar declines in epiretinal membranes and retinal vein occlusion beyond standard visual acuity. Across all disease states, color and contrast vision were negatively impacted.

## Introduction

Human trichromatic vision enables the detection of 2.3 million colors with discrimination between wavelength differences of as little as 0.25 nm.^1^ Despite 50 percent of visual information originating from color-sensitive cones in the fovea, clinical visual function testing is largely limited to visual acuity (VA) which quantifies minimal angle of resolution under maximal contrast (black-on-white) conditions. With its ease of use and wide accessibility, VA is the most used visual function test and is the “gold standard” for retinal disease clinical trials. Techniques have been developed to mitigate visual acuity charts test results,^2^ including the Early Treatment of Diabetic Retinopathy Study (ETDRS).^3^ However, the contributions of color and contrast to visual function are merely noted subjectively in patients with ophthalmic disease. Routine color vision deficit (CVD) testing in practice is typically reserved for neurophthalmic disease, orbital compressive pathologies, and hereditary reitnopathies^4-6^ with advantages and disadvantages for the various color and contrast assays.^1^ The anomaloscope, considered the “gold standard” quantitative matching test for assessing CVD,^1,7^ is rarely used because it is laborious, time-consuming, and restricted to quantifying CVD in the red-green axis. Instead, pseudoisochromatic plates are frequently used because they are easy to administer, rapid, and inexpensive; however, they provide low resolution quantitative results.^1,7^ Quantitative arrangement tests like the Farnsworth Munsell 100^8^ and computerized adaptations are lengthy and performance is confounded by external factors including nonverbal intelligence.^9^

The ColorDx Cone Contrast Threshold Test (CCT) (ColorDx HD, Konan Medical, Irvine, CA), a computerized CVD test developed for the US Air Force, uses an adaptive algorithm to consecutively evaluate contrast sensitivity in L-, M-, and S-cones.^10^ Individuals are presented with a series of tumbling Landolt-C optotypes in a randomized orientation of cone contrast against an isochromatic photopic (∼74 cd/m2) background (**Fig. 1a**) and prompted to indicate the orientation within 5 seconds. ColorDx generates a continuous quantitative output for each of the 3 cone opsins (**Fig. 1a**). This test is 4-8 minutes long, is easy to administer, and can be standardized between testing centers with pre-programed color and luminance calibration on an anti-glare screen. Military screenings using CCT demonstrate 96-100% sensitivity and specificity for protan, deutan, and tritan CVD compared to the anomaloscope.^1,11^ Our study team has also demonstrated in data under review that there was no learning effect, or improvement in scores, when individuals repeated this test multiple times^12^

**Figure 1:**
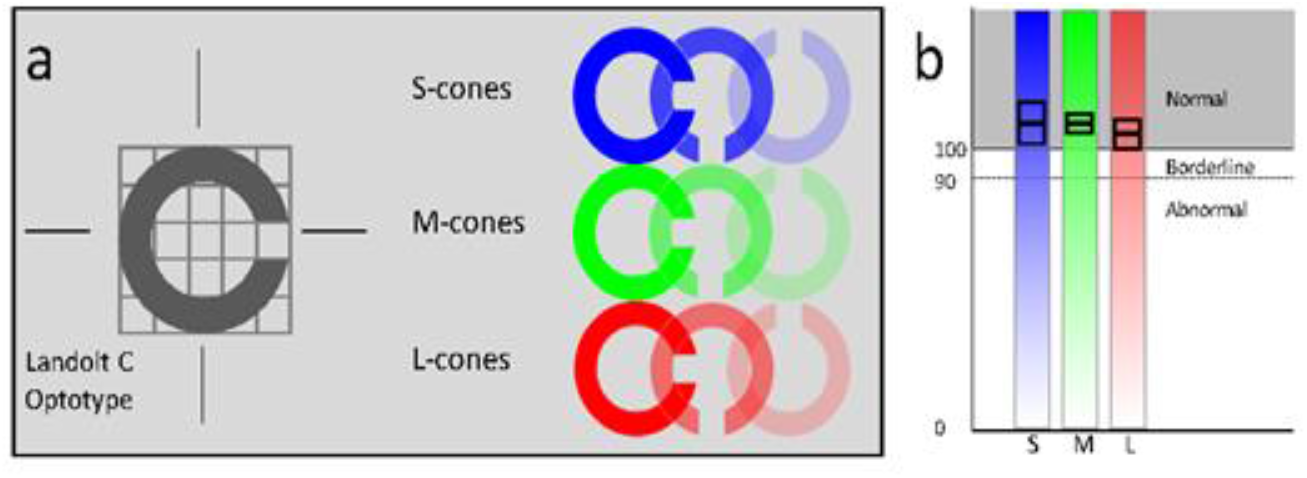
**a)** Cone contrast threshold test isolates cone opsins by providing photopic background to suppress rod function and adjusts Landolt C optotype contrast for each cone opsin. **b)** The quantitative output indicates a mean score with error for each individual cone opsin and these are reported as normal, borderline or abnormal.

We report visual function changes beyond visual acuity in patients with 4 diseases affecting the neurosensory retina. Loss of retinal ganglion cells with or without^13^ optic neuritis (ON) in multiple sclerosis (MS) has established serial color vision testing as standard care for optic neuropathies. Color and contrast vision is not standard care in diseases affecting the retina. Age-related macular degeneration (AMD) is a common cause of vision loss primarily metabolic dysfunction^14^ and death^15^ of the photoreceptors and retinal pigmented epithelium, and CCT has been shown be reduced in the intermediate stage of the disease.^16^ Epiretinal membranes (ERM) are thin fibrocellular membranes which exert mechanical force from the inner retina to the outer retina to reduce visual quality and acuity, but little is known about their effect on color and contrast vision.^17^ Visual acuity loss from retinal edema associated with **retinal vein occlusion** (RVO), a vascular disease of the inner retina, is well treated with injectable medications;^18^ however, patients still note poor vision despite good visual acuity recovery. In AMD, ERM, and RVO, patients subjectively report qualitative reductions in vision despite normal visual acuity (see images, Supplemental Digital Content 1-3, with clinical images and corresponding CCT for such examples). The aim of this study is to utilize CCT quantitative cone contrast vision testing in patients with MS, AMD, ERM, and RVO to characterize visual function deficits beyond BCVA.

## Methods

### Human Subjects

We performed a retrospective chart review on 268 patients seen at the Gavin Herbert Eye Institute during the period of May 2018 through December 2019. Institutional Review Board (IRB)/Ethics Committee approval was obtained, and the described research adhered to the tenets of the Declaration of Helsinki. We included patients if they had one of several diagnoses including MS with and without previous ON, AMD, ERM, or RVO. ColorDx is not a visual acuity test but requires 20/120 vision or better to complete the test. All subjects across the various disease states had to meet the following criteria: age 18 or older, no history of other ocular comorbidities except for age-related cataracts, VA of 20/60 or better, and no prior history of ocular surgery except for cataract extraction. MS patients were compared to an age and visual acuity-matched healthy control group subject to the same inclusion and exclusion criteria. All subjects had previously obtained CCT and VA assessments (Snellen letter set, Sloan characters with ETDRS spacing as is customary on M&S Technologies digital visual acuity charts) as a part of routine care at the Gavin Herbert Eye Institute.

Retina specialists made the diagnosis of AMD, ERM, and RVO (AB, MM). Neurologists and neuro-ophthalmologists made the diagnosis of MS (CB, WC). Specialists performed dilated retinal exam and optical coherence tomography (OCT) to diagnose and grade AMD by evaluating the extent of pigmentary changes, drusen number and drusen size. ^19,20^ Dilated retinal exam and SD-OCT imaging confirmed the presence of ERM. Two independent retina specialists determined severity and grading^17^ of ERM (AB, DF). Patients were excluded if they were actively receiving treatment for AMD, defined as having received any injections of steroid or anti-VegF within 3 months of performing the CCT assessment. Eyes affected by ERM and AMD were compared to an age and visual acuity-matched subset of fellow healthy eyes meeting the following criteria: trace or no ERM and drusen or no sign of early AMD. Diagnosis of RVO was based on clinical examination and a historical onset of more than 6 months and no active macular edema after treatment.

### Cone contrast threshold and imaging

ColorDx is a CCT test that presents a Landolt C optotype in 4 different orientations in the center of a photopic gray screen. Patients use a response pad with arrows to indicate optotype orientation. Following color and luminance calibration on an anti-glare screen, the device presented tumbling Landolt-C optotypes. A Bayesian thresholding method, the Psi-Marginal Adaptive Technique, adjusted the contrast for subsequent tests based on preceding answers^10^ and a Psi-marginal calculation yielded the final CCT value.

The device utilizes an adaptive algorithm per manufacturer specifications to increase or decrease the contrast of the displayed optotype in accordance to the test subject’s response accuracy. All CCT testing was performed with BCVA under photopic and monocular conditions at 2 feet. BCVA, age, and phakic status were recorded for all subjects. SD-OCT (Cirrus, Carl Zeiss Meditiec, USA or Spectralis Heidelberg Engineering Heidelberg Germany) imaging was acquired for all subjects in the disease state groups.

### Statistical Analysis

As each patient’s eyes were subject to different retinal conditions, we considered each eye a separate unit of analysis. Sample characteristics (mean, standard deviation and range) with respect to age and visual acuity (logMAR) were calculated for all eyes across groups. Then we performed linear regression models to estimate the mean differences in CCT scores for each cone class by ERM grade, AMD severity, and, among MS patients, by diagnosis of active optic neuritis and total RNFL thickness (G) or temporal RNFL octant thickness (T) each compared to healthy controls as the reference group. To assess RVO status, we used linear regression models to compare affected vs. unaffected eye in each patient. All models were estimated using generalized estimating equations (GEE) to account for clustering within persons and adjusted for age, visual acuity and phakia. For analyses by ERM grade and AMD severity, we chose healthy controls restricted to the age range of the disease population. For each analysis, individual CCT score distributions for each cone class were visually compared using violin plots. with overlying mean CCT score differences with 95% confidence intervals (CIs) for each group. Comparisons for significance were made with healthy controls using adjusted regression model estimates. We conducted all analyses using R version 1.2.^21^

## Results

**Table 1** summarizes the distribution of age and visual acuity all subjects. CCT results are represented in violin plots, with distributions of patients on the horizontal axis with the group CCT scores plotted from 0 to 150 on the vertical axis. The adjusted mean and 95% confidence interval for each plot are overlaid. Significant differences from healthy controls are represented by asterisks.

### Multiple Sclerosis

Not all patients with MS were able to complete binocular CCT as some patients had monocular vision loss which precluded test performance. MS patients with a history (9 eyes) and without a history (22 eyes) of ON demonstrated reduced CCT scores compared to the healthy control subject group. Likewise, MS was associated with significant reductions in CCT scores for all cone classes even among those with normal RNFL, except for L-opsin in which normal RNFL was not associated with a significant reduction in CCT (**Fig. 2**). There was a downtrend in CCT performance in eyes with abnormal RNFL compared to normal RNFL across all 3 color cones. There were no statistically significant correlations between VA and CCT score or VA and RNFL.

**Figure 2:**
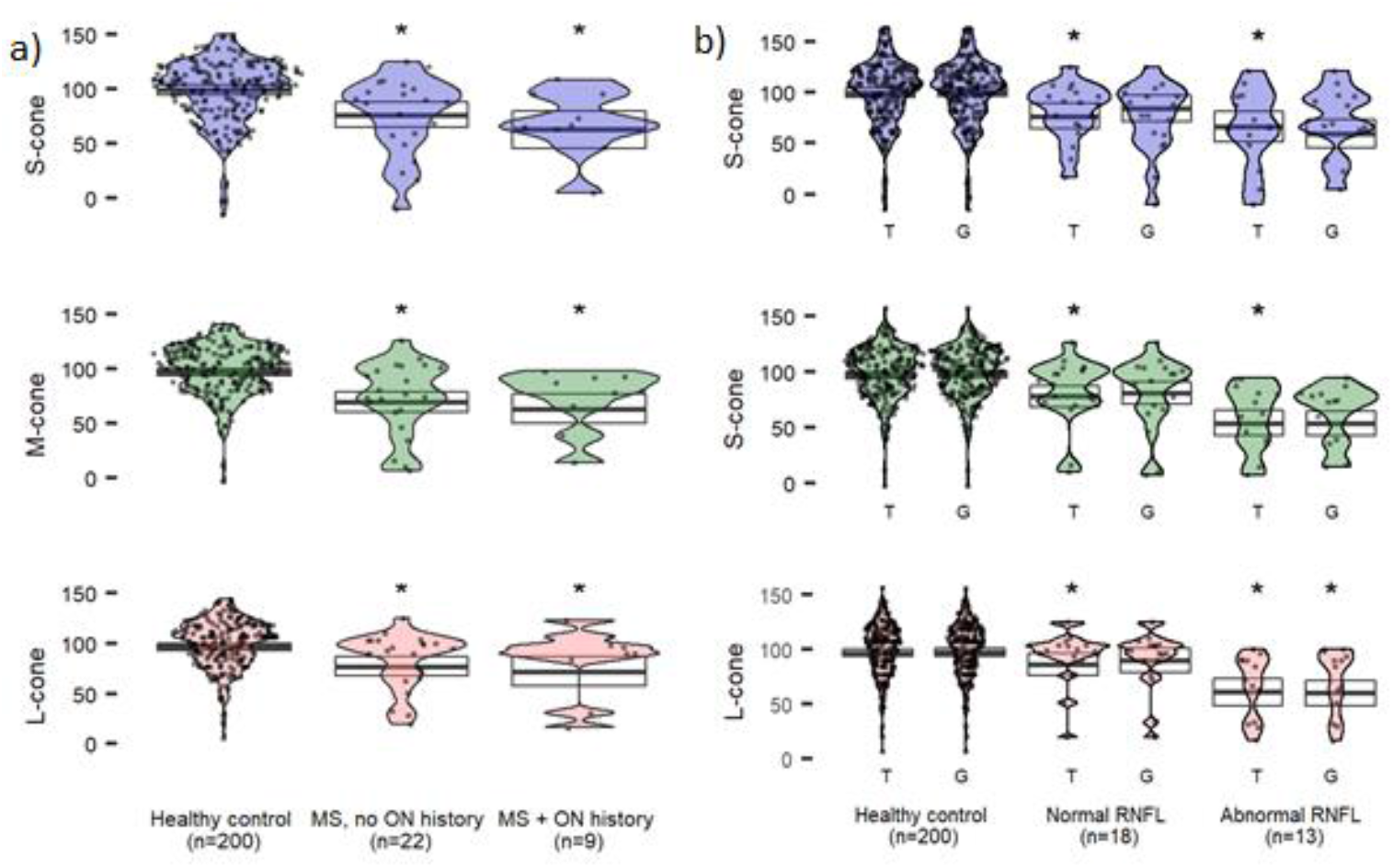
CCT in patients with MS compared to healthy controls. **a**) CCT scores in eyes with and without a history of optic neuritis. **b**) CCT scores in patients with RNFL thinning in the temporal (T) octant, or the average (G) of all octants. ON = Optic neuritis, RNFL = retinal nerve fiber layer. Estimated means (95% CI) are adjusted for age, visual acuity and phakic status. Asterisks indicate significant differences (vs healthy) at p<0.05.

### Age-related Macular Degeneration

All classification stages for AMD, except early AMD, demonstrated significant (p < 0.05) reduction in CCT scores (**Fig. 3**). Advanced AMD with or without neovascularization represents the largest reduction in CCT scores demonstrated by widening of the lowest portions of the violin plots. Additionally, no subjects had normal M-cone and L-cone CCT with rare patients in the neovascular AMD retaining some tritan vision (s-cone plot is taller for neovascular AMD than advanced non-neovascular AMD).

**Figure 3:**
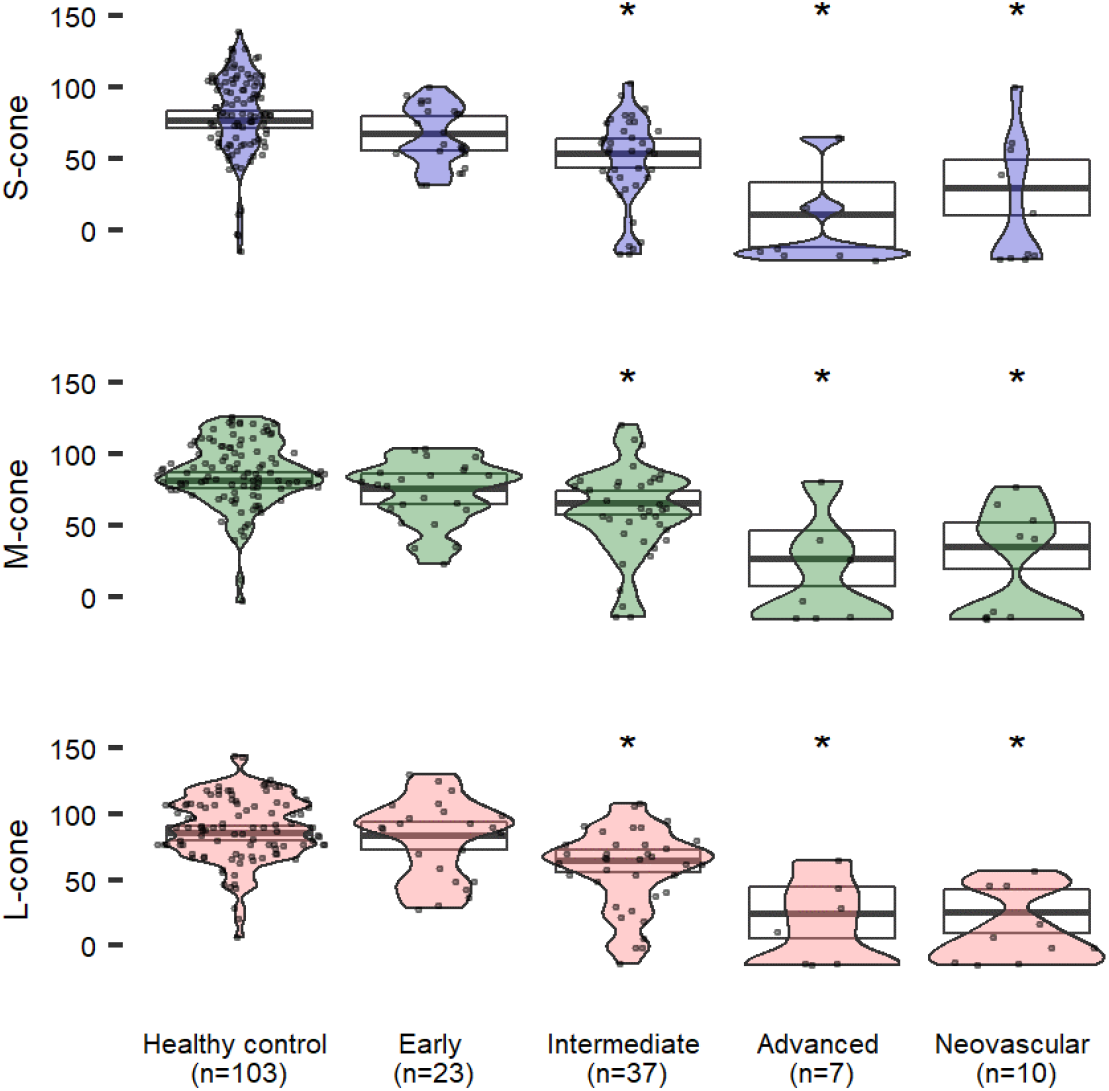
CCT scores in patients with age related macular degeneration at different stages compared to healthy controls ≥50 years old (matched to age range of AMD patients). Estimated means (95% CI) are adjusted for age, visual acuity and phakic status. Asterisks indicate significant differences (versus healthy) at p<0.05.

### Epiretinal Membranes

Two retina specialists classified subjects diagnosed on clinical exam and SD-OCT into ERM grades. CCT results are reported in **figure 4**. Each grade of ERM demonstrated a statistically significant reduction in CCT compared with healthy controls (p< 0.05). Both S- and M-cones demonstrate a nominal reduction in CCT with advancing ERM grade which is not observed with the L-cone. For all ERM grades, groups of patients with normal CCT scores are seen in the upper half of each violin plot, while a broadening of the lower half of the plot is seen for each ERM grade.

**Figure 4:**
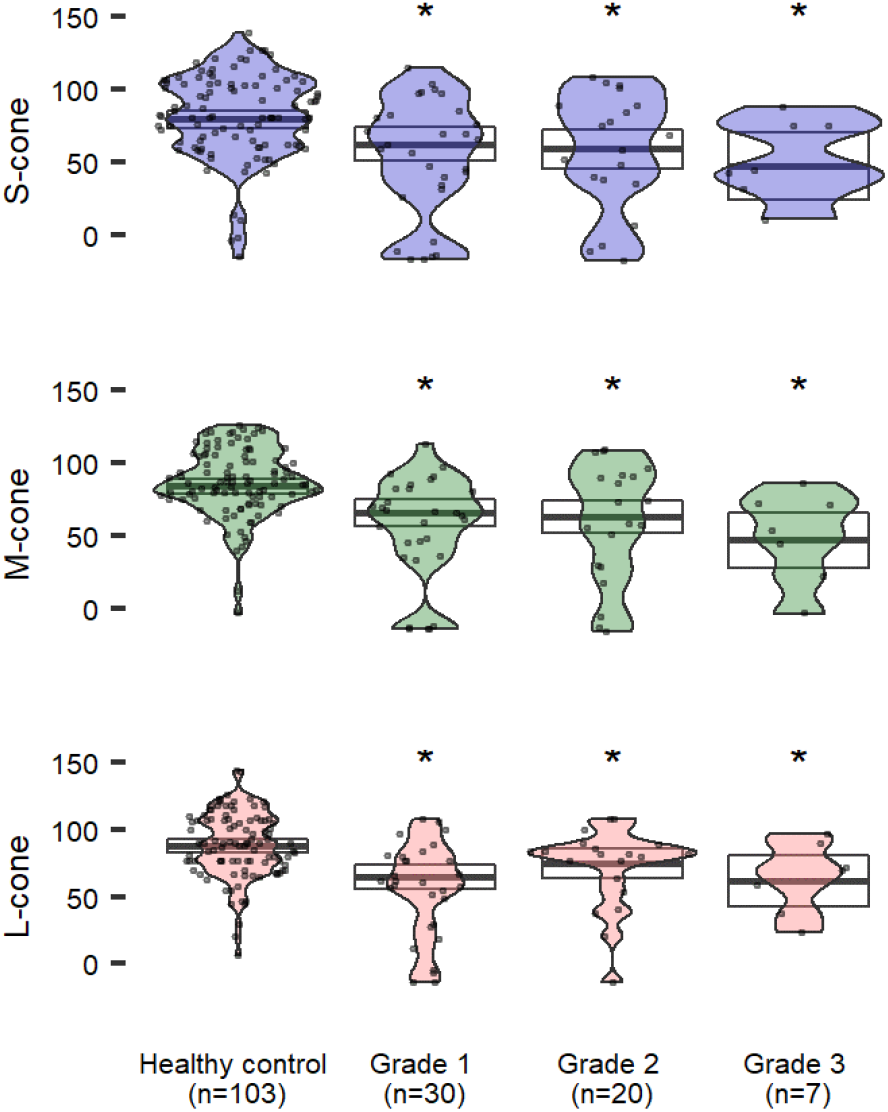
CCT scores in patients with epiretinal membranes increasing in severity compared to healthy controls ≥50 years old (matched to age range of ERM patients). Estimated means (95% CI) are adjusted for age, visual acuity and phakic status. Asterisks indicate significant differences (versus healthy) at p<0.05.

### Retinal Vein Occlusion

All patients with RVO (n=11) who were evaluated with CCT in this study had achieved steady state resolution of macular edema and had sufficient visual acuity scores to reliably complete CCT testing (ranging 20/25 to 20/100). All eleven patients (86%) had visual acuity of 20/60 or better (twice the minimum angle of resolution to reliably complete CCT testing). There was no significant correlation observed between visual acuity and CCT (**Fig. 5**). For example, one patient with 20/100 visual acuity had near zero CCT results which were similar to other eyes with 20/25 visual acuity. Another patient with 20/80 visual acuity had diminished, but not zero, CCT scores equivalent to other eyes affected by RVO and 20/25 vision.

**Figure 5:**
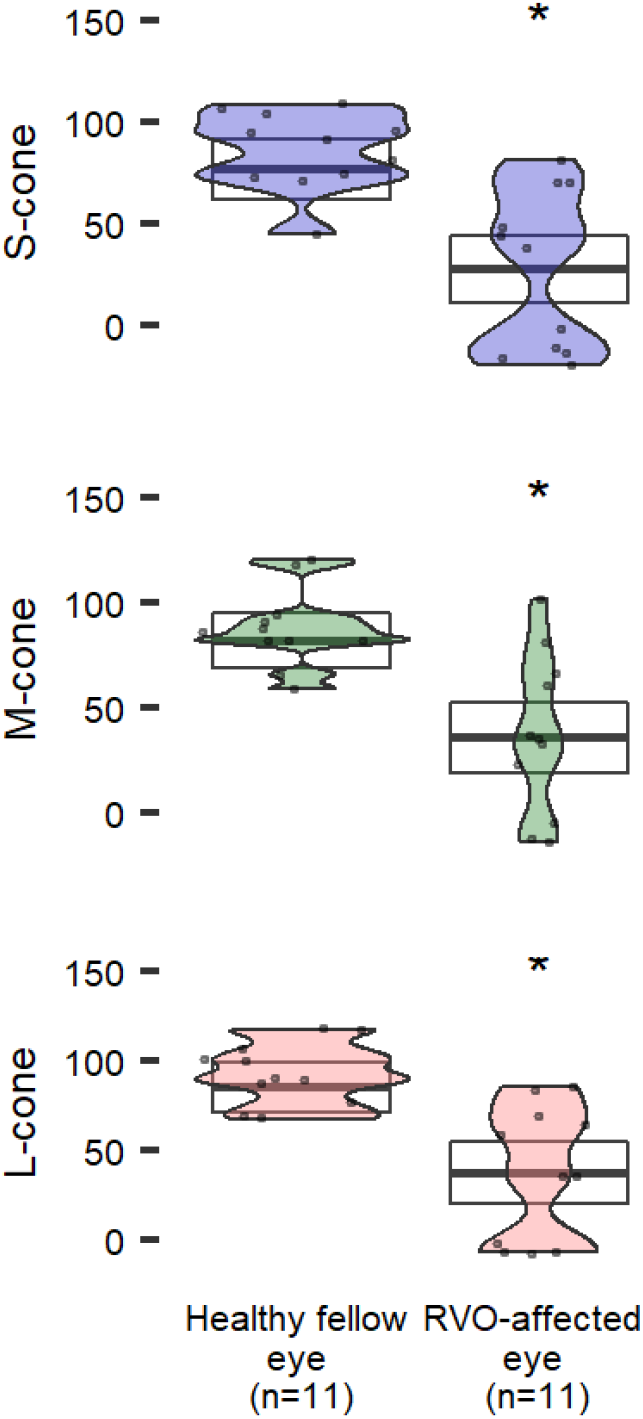
CCT in patients with retinal vein occlusion. Estimated means (95% CI) are adjusted for age, visual acuity and phakic status. Asterisks indicate significant differences (vs healthy) at p<0.05.

## Discussion

We analyzed cone contrast threshold testing of patients with a variety of diseases with differing pathophysiological mechanisms for vision loss. CCT performance has been shown to quantify contrast and color vision. For these various diseases, we sought to determine if CCT reveals diminished visual function beyond VA alone. We first verified color vision deficits in a disease for which color vision testing is performed as standard of care, MS. We then evaluated CCT testing in retinal diseases for which color and contrast vision is not routinely tested.

**Multiple Sclerosis** is associated with earlier and more pronounced color and contrast vision dysfunction compared to other eye diseases.^22^ Color vision loss in MS was previously thought to occur secondary to ON, due to direct damage to the optic nerve and resulting damage to the retinal ganglion cell layer.^6^ More recently, retinal fiber nerve layer atrophy and reduced macular volume have been described in MS patients with no history of ON.^13^ One theory suggests a secondary retinopathy in MS due to retrograde axonal degeneration.^13^

Our study demonstrated that MS with no history of ON exhibited a significant reduction in CCT compared to controls (**Fig. 2a**). This supports that functional vision loss occurs as a result of a separate disease process from optic nerve inflammation. The downtrend in CCT scores with abnormal RNFL compared to normal RNFL implies suggests a correlation between functional vision loss and RNFL although a larger sample size is needed to further correlate this relationship (**Fig. 2b**).

The temporal octant of the optic nerve transmits visual signals from the fovea and perifovea. Thinning of the temporal octant is associated with a significant reduction in CCT scores, and the same is true for the global nerve fiber layer thickness in the optic nerve. Even after adjusting for visual acuity, visual quality described by CCT performance declines. Because global and temporal changes in RNFL thickness correlate with CCT reduction, but only the temporal RNFL serves CCT performance, changes in analogous visual quality features may continue in the eccentric and peripheral vision.

**Age-related macular degeneration** is the leading cause of irreversible blindness in people older than 50 years, the leading cause of visual disability in the industrialized world and the third leading cause of visual disability globally.^23^ Drusenoid deposits with progressive RPE and outer retinal atrophy are hallmarks for the disease.^24^ Dysfunctional processing of lipid metabolites, photoreactive retinoids, and aberrant accumulation of bisretinoids initiate drusen formation beneath the retina and retinal pigment epithelium. A consequence of cellular dysfunction, the physical presence of drusen further disrupts normal retinal anatomy, physiology and vision. Early AMD is functionally indistinguishable from normal aging;^16^ however, intermediate disease produces reduced color and contrast sensitivity with poor low-light vision which can precede the onset of visual acuity loss. Advanced AMD extinguishes central visual acuity and contrast vision. Clinical trials directed at treating or slowing progression of diseases like non-neovascular (dry) AMD rely exclusively on subjective VA determinations and objective structural imaging. Some new treatments show promise because they slow structural damage^25^ while other potential treatments are discarded after multimillion dollar clinical trials because they do not show the same structural benefits.^26^ While retinal structure and visual acuity do correlate, subjective measures of visual function extend beyond visual acuity to contrast sensitivity, color perception, and the ability to respond to changing lighting conditions. These other subjective visual function tests are not routinely performed because they are slow, require trained technical administration, demand reliable patient participation, and do not integrate easily with a busy clinic schedule.

AMD patients described in this report confirm similar findings to Cocce et. al.^22^ with CCT results insignificantly changed in early AMD, but intermediate and advanced AMD yield large reductions in color vision. CCT may be used as a clinical endpoint for interventions directed at intermediate AMD and beyond. Inability for conventional testing to identify functional endpoints for early AMD highlights our communities need for novel assays sensitive to early disease.

**Epiretinal membranes** are thin avascular fibrocellular membranes which develop on the inner surface of the retina. ERMs can be asymptomatic or alter macular structure to produce metamorphopsia and reduced visual acuity. SD-OCT studies of the ERM reveal retinal disruptions that may impair cone function such as blurred, interrupted, or absent cone outer segment tip lines (COST).^27^ Additionally, the ectopic inner foveal layers (EIFLs) within ERMs have been associated with significant visual function loss.^17^ Disruptions of the COST line has been associated with macular diseases and may indicate photoreceptor dysfunction.^28^ Recovery of the COST after ERM surgery correlates with better outcomes in VA, with COST thickness prior to ERM surgery correlating well to postoperative VA.^29^ While some patients subjectively report metamorphopsia and reduced acuity, others report reduced quality in their vision (see image, Supplemental Digital Content 2, example ERM clinical images and corresponding CCT). However, little is known about the qualitative changes in vision beyond visual acuity.

Utilizing CCT, we demonstrate significant color and contrast deficits in patients with ERM’s of mild to severe grade. While not statistically significant, there was a downtrend in CCT performance with worsening ERM grade in L and M cones and further research is warranted to determine if disease severity correlates with the degree of color and contrast vision loss.

**Retinal vein occlusion** remains one of the most investigated retinal disease conditions because intraocular anti-VEGF and steroid injections demonstrate profound improvements in retinal anatomy seen on OCT and visual acuity.^30^ RVO, like diabetic retinopathy, is an inner retinal vascular disease affecting cells in the ganglion cell layer and inner nuclear layer. Clinical trials to treat eyes affected by RVO focus on visual acuity and OCT anatomy as the primary outcome for therapeutic success. However, patients who have recovered visual acuity suitable for reading or driving remain dissatisfied with the quality of their vision (see image, Supplemental Digital Content 3, example of RVO clinical images and corresponding CCT).

CCT testing in patients who have recovered optimal visual acuity and retinal anatomy highlights the residual loss of color and contrast vision in eyes affected by RVO when compared with unaffected fellow eyes. This finding was present for both eyes with 20/40 vision or better (n=8), and eyes with vision worse than 20/40 (n=6). CCT, therefore, presents a practical quantitative endpoint for therapies aimed at optimizing visual quality in addition to visual acuity.

Results of this study are likely impacted by its retrospective nature, and limited sample size, particularly for the higher grade ERM states. ETDRS testing was not used because all data was collected during standard course of care where the M&S technologies digital display for visual acuity testing. While this report highlights changes in color and contrast vision with disease, further prospective studies with larger sample sizes are warranted.

## Conclusion

As one of the first studies to utilize CCT testing in MS, AMD, ERM, and RVO patients, we present novel quantitative data describing cone-specific visual function across all 3 cone classes in these disease states. We observed that inner retinal vascular and inner retinal mechanical disease states both diminish CCT test results, suggesting that CCT performance is affected by multiple mechanisms downstream of photoreceptors.

## Data Availability

Available upon request

## Acknowledgments

The authors would like to thank Donald Frambach MD for assistance with grading ERM and Konan medical for an unrestricted donation of ColorDX CCT devices.

---

**Supplementary Figure 1:**
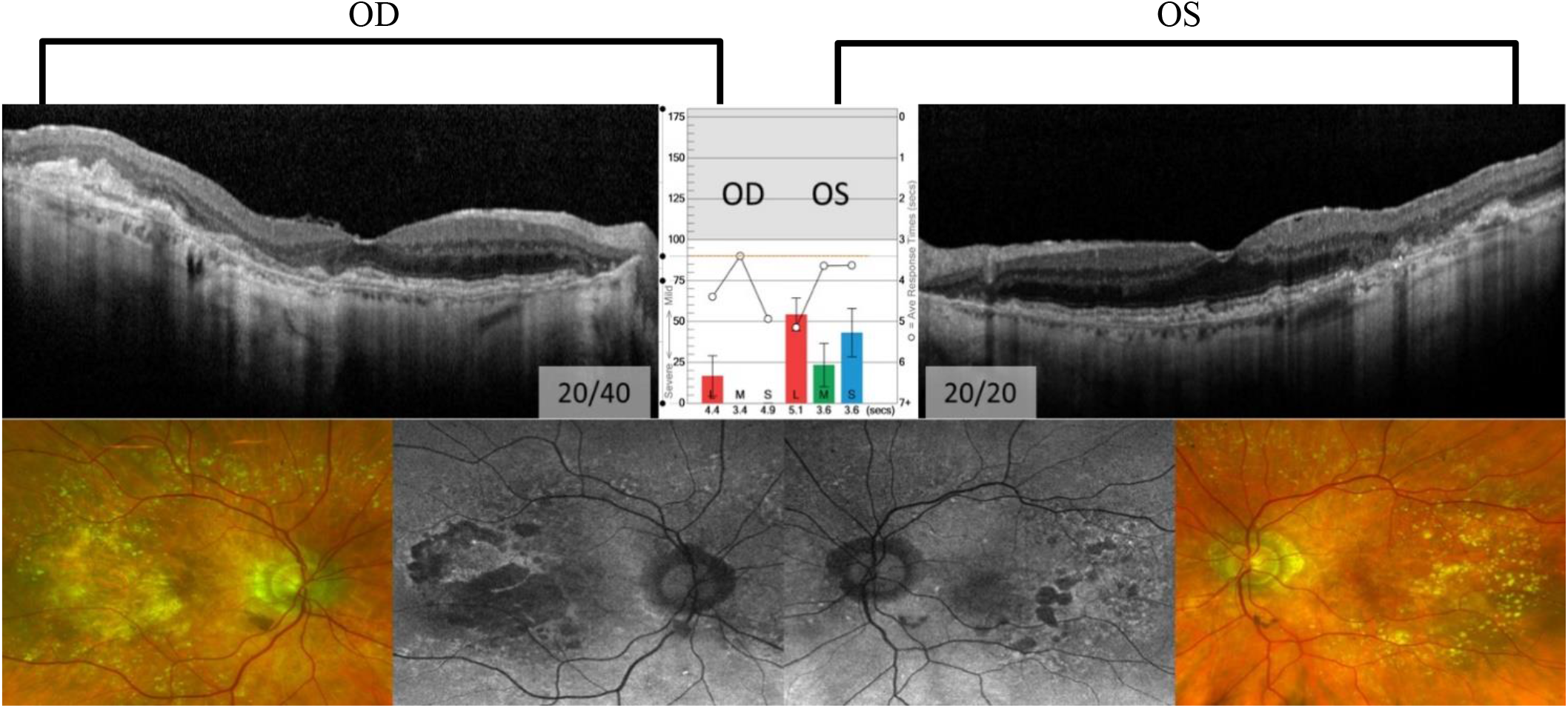
94-year-old pseudophakic male with quiescent nvAMD in his right eye and nnvAMD in his left eye. The patient reports that color blocks in the CCT report appear black in his right eye (VA: 20/40), and washed out in his 20/20 left eye (VA: 20/20).

**Supplementary Figure 2:**
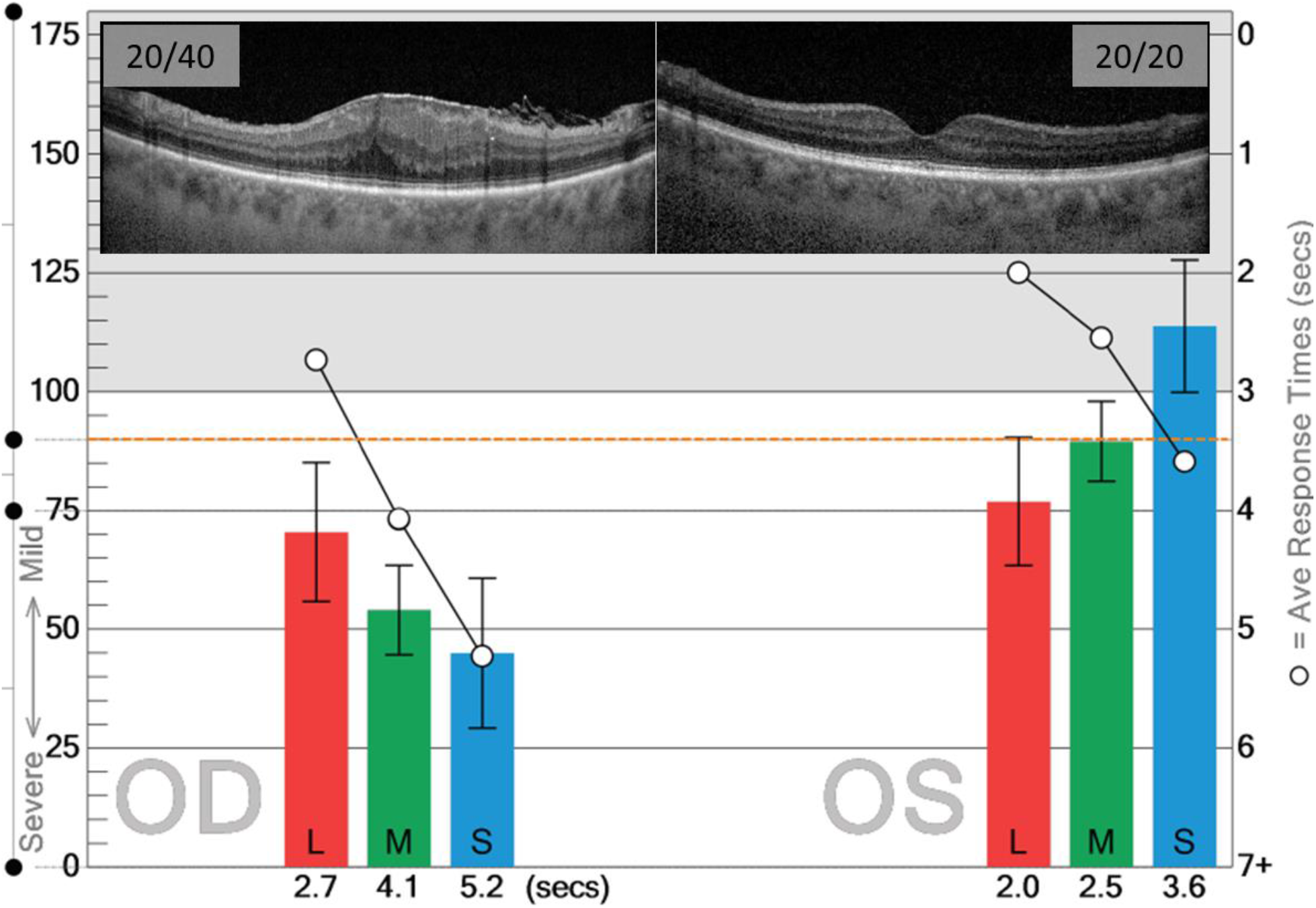
72-year-old pseudophakic female reports visual distortion and 20/40 vision OD with normal 20/20 vision OS. SD-OCT imaging of right eye demonstrates ERM with corresponding reduction in CCT. *Orange line denotes cut-off point for normal cone contrast scores.

**Supplementary Figure 3:**
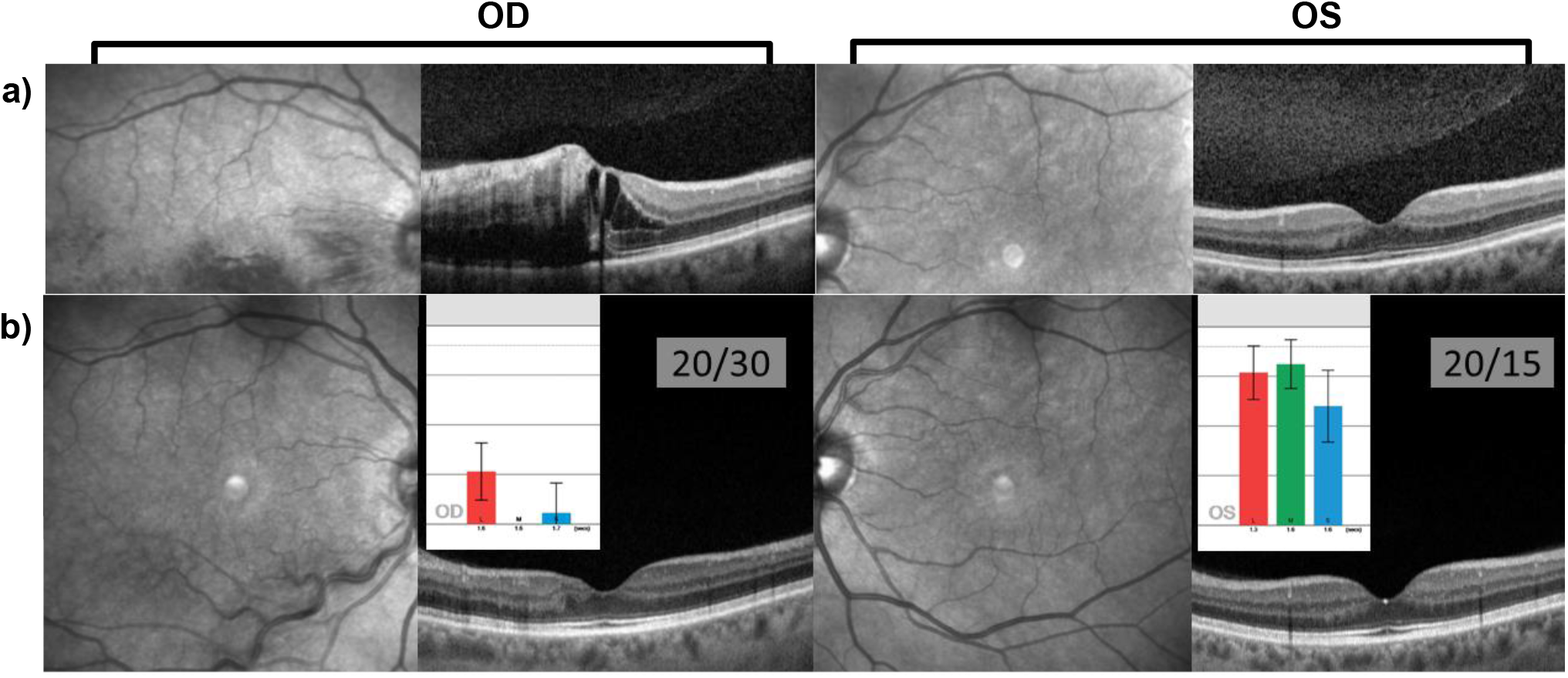
**a)** Fundus Autofluorescence images and visual acuities for 65-year-old phakic female who presented with acute onset blurry vision OD was found to have an inferior hemiretinal RVO and cystoid macular edema. **b)** After anti-VEGF therapy the patient’s vision improved from 20/60 to 20/30 but she remained very symptomatic for “poor” vision. CCT scores OD were significantly lower and near zero OD compared to OS even when visual acuity was 20/30 and 20/15.

## Notes

**Financial Support:** This project was supported by the National Center for Research Resources and the National Center for Advancing Translational Sciences, National Institutes of Health, Bethesda, MD, through RPB unrestricted grant to UCI Department of Ophthalmology ICTS KL2, grant numbers: KL2 TR001416 and UL1 TR001414

### Competing Interest Statement

The authors have declared no competing interest.

### Funding Statement

This project was supported by the National Center for Research Resources and the National Center for Advancing Translational Sciences, National Institutes of Health, Bethesda, MD, through RPB unrestricted grant to UCI Department of Ophthalmology
ICTS KL2, grant numbers: KL2 TR001416 and UL1 TR001414

### Author Declarations

University of California Irvine IRB aproval HS#2019-5254 and HS#2019-5411

